# Comparison of the risk of hospitalisation among BA.1 and BA.2 COVID-19 cases treated with Sotrovimab in the community in England

**DOI:** 10.1101/2022.10.21.22281171

**Authors:** Katie Harman, Sophie G Nash, Harriet H Webster, Natalie Groves, Jo Hardstaff, Jessica Bridgen, Paula B Blomquist, Russell Hope, Efejiro Ashano, Richard Myers, Sakib Rokadiya, Susan Hopkins, Colin S Brown, Meera Chand, Gavin Dabrera, Simon Thelwall

## Abstract

**Objectives:** Sotrovimab is one of several therapeutic agents that have been licensed to treat people at risk of severe outcomes following COVID-19 infection. However, there are concerns that it has reduced efficacy to treat people with the BA.2 sub-lineage of the Omicron (B.1.1.529) SARS-CoV-2 variant. We compared individuals with the BA.1 or BA.2 sub-lineage of the Omicron variant treated Sotrovimab in the community to assess their risk of hospital admission.

**Methods:** We performed a retrospective cohort study of individuals treated with Sotrovimab in the community and either had BA.1 or BA.2 variant classification.

**Results:** Using a Stratified Cox regression model it was estimated that the hazard ratios (HR) of hospital admission with a length of stay of two or more days was 1.17 for BA.2 compared to BA.1 (95% CI 0.74-1.86) and for such admissions where COVID-19 ICD-10 codes was recorded the HR was 0.98 (95% CI 0.58-1.65).

**Conclusion:** These results suggest that the risk of hospital admission is similar between BA.1 and BA.2 cases treated with Sotrovimab in the community.

## Background

The use of therapeutic agents for the treatment of COVID-19 is a rapidly evolving field of research where several new and existing treatments have been licensed to help treat COVID-19 infection in people who are at highest risk of developing severe outcomes. In the United Kingdom (UK), neutralising monoclonal antibody (nMAb) Sotrovimab was approved in December 2021 for use in symptomatic people who do not require oxygen but are at increased risk of severe infection. It was licenced to treat SARS-CoV-2 infection both for people in the community and those hospitalised (1).

Initial analyses of Sotrovimab have shown that its administration significantly reduced the risk of hospitalisation or death in people with mild to moderate infection who had at least one risk factor for developing severe outcomes (2, 3). However, findings from in vitro studies have raised concerns that Sotrovimab may have reduced clinical efficacy for people with the BA.2 sub-lineage of the Omicron (B.1.1.529) variant due to mutations in the spike protein causing reduced neutralising ability (4, 5). These concerns led to withdrawal of Sotrovimab authorisation in several countries including the United States of America (6). Sotrovimab is still licenced for use in the UK but it is currently the third-line recommended treatment for people in hospital (7) and as of June 2022 is more commonly given to people in the community (8). An initial analysis by the UK Health Security Agency (UKHSA) found that there was no increased risk of attendance or admission to hospital, however this was limited by reduced follow up time (9). Therefore, this study sought to assess whether there were any differences in hospital admission among people who were treated with Sotrovimab in community settings with BA.2 compared to those with BA.1 sub-lineages.

## Methods

Information about individuals treated with Sotrovimab in the community submitted by NHS Trusts up to 24 May 2022 were extracted from Blueteq software and linked deterministically by NHS number to laboratory confirmed COVID-19 cases in England from UKHSA’s Second Generation Surveillance System (SGSS). This incorporates data on variant status from genotyping, S-gene target data and Whole Genome Sequencing (WGS) (10). Vaccination status of these cases was then obtained by linking to the National Immunisation Management Service (NIMS), and categorised based on receipt of vaccine at least two weeks prior to positive specimen date. Cases were then linked to NHS Digital’s Emergency Care Data Set (ECDS) and Secondary Uses Service (SUS), for information about their hospital admissions as described elsewhere (11).

Genotyping can identify the Omicron variant but is unable to distinguish sub-lineages. However, the S-gene target status can be used in conjunction with genotyping PCR results to identify BA.1 and BA.2 for samples tested with the TaqPath assay, and therefore was used in conjunction with genotyping results, or used solely when Omicron was the dominant variant in circulation. The study therefore included: (1) all sequencing-confirmed BA.1 and BA.2 cases; (2) between 1 January and 24 January 2022 when Delta (B.1.617.2) variant was prevalent, SGTF/SGTP information was only used if genotyping PCR results indicated Omicron; and (3) from 25 January to 3 May 2022 all SGTF/SGTP cases were included irrespective of genotype status as predictive values of SGTF/SGTP to call BA.1/BA.2 were ≥95% (12).

Hazard ratios (HR) of hospitalisation outcomes for those treated with Sotrovimab with the BA.2 variant compared to BA.1 were estimated using Stratified Cox regression models. Hospital admission was defined as: (1) hospital admission with either a length of stay of two or more days, or the person died in hospital, (2) hospital admission with either a length of stay of two or more days, or the person died in hospital, and mention of COVID-19 ICD-10 codes (U071/U072). All definitions were restricted to be within 0-14 days from the date of treatment and excluded hospital admissions for injury-related reasons. Models were stratified by specimen week and then adjusted for age group, linear effect in age and vaccination status, to account for confounders. All analyses were performed in R (version 4.1.2, R Foundation for Statistical Computing, Vienna, Austria).

## Results

Between 01 January and 26 April 2022 20,274 records for community prescription of Sotrovimab were received, 19,149 (94.5%) of which successfully linked to the COVID-19 case episodes list. 8,850 (46.2%) had BA.1 or BA.2 variant classification data available and were therefore included in analysis, 4,285 for BA.1 and 4,565 for BA.2 respectively (Table 1).

**Table 1:**
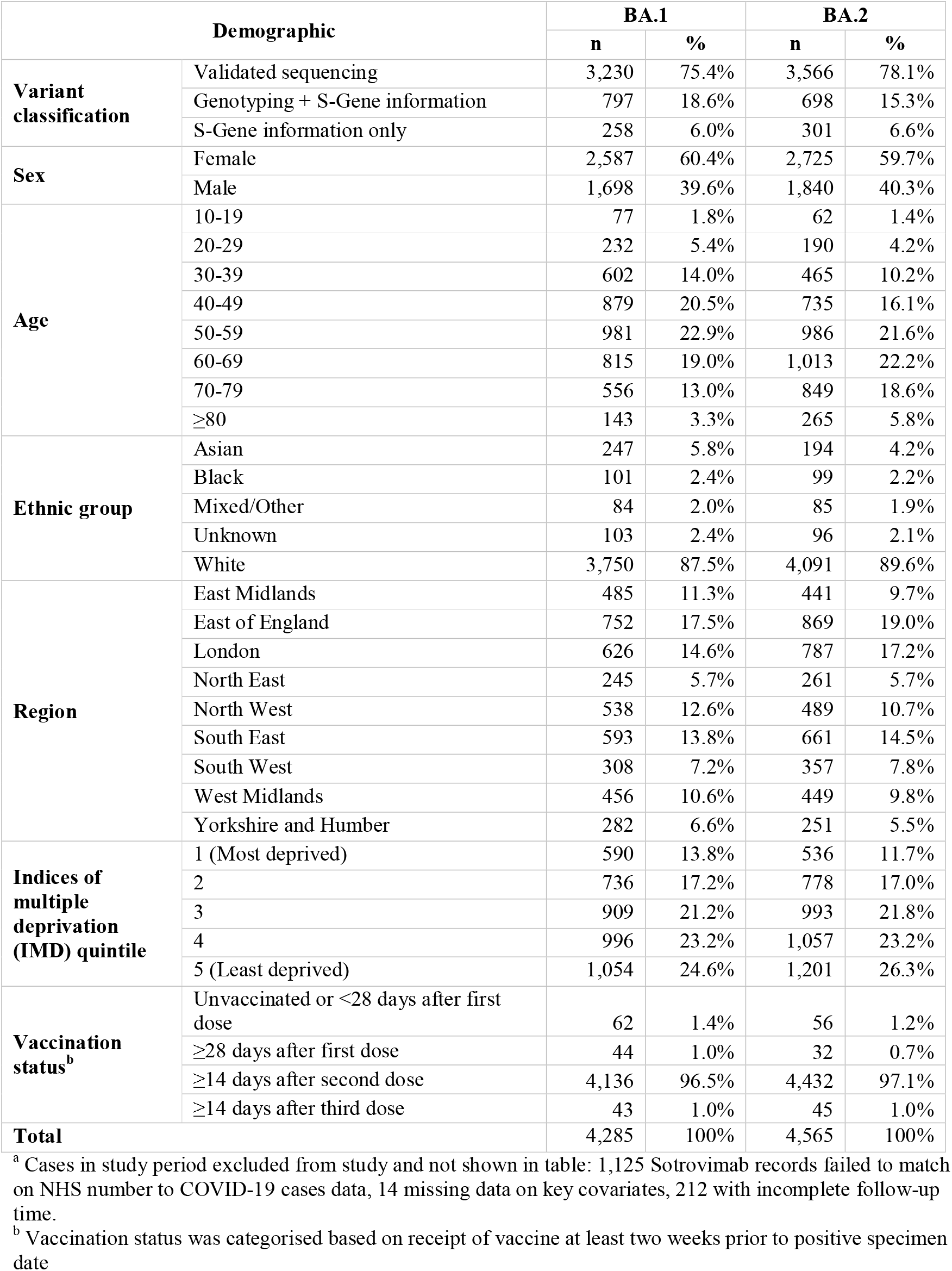
Demographic characteristics of COVID-19 cases treated with Sotrovimab in the community with Omicron lineage BA.1 or BA.2: England, 1 January to 26 April 2022 (n = 8,850^a^)

The patient characteristics were broadly similar between the two cohorts, with the same proportions by sex. The BA.1 cohort had a younger age distribution compared to BA.2, where the median age of individuals with BA.1 was 53 years (interquartile range 41-64) compared to 58 years for BA.2 (interquartile range 46-69). The geographical breakdown was broadly similar between the two groups, however slightly more of the BA.2 cohort resided in London compared to the BA.1 cohort (p<0.001), while a slightly larger proportion of the BA.1 cohort resided in more-deprived areas compared to the BA.2 (p<0.001).

The risk of hospital admission with a length of stay of 2 or more days within 14 days of community treatment with Sotrovimab showed no statistically significant difference between BA.2 and BA.1 sub-lineages (HR=1.17, 95% CI 0.74-1.86; Table 2). A sensitivity analysis restricting to hospital records which stated COVID-19 ICD-10 codes on admission for greater specificity also showed no statistically significant difference in hospitalisation risk in BA.2 cases compared to BA.1 (HR=0.98, 95% CI 0.58-1.65).

**Table 2:** Risk of hospitalisation for COVID-19 cases treated with Sotrovimab in the community with Omicron lineage BA.1 or BA.2: England, 1 January to 26 April 2022

| Outcome | Count of outcome in BA.1 cases | Count of outcome in BA.2 cases | Adjusted hazard ratio (95% CI)<sup>a</sup> |
| --- | --- | --- | --- |
| Hospital admission with length of stay ≥ 2 days | 91 | 77 | 1.17 (0.74 - 1.86) |
| Hospital admission with length of stay ≥ 2 days and mention of COVID-19 ICD-10 codes | 73 | 62 | 0.98 (0.58 - 1.65) |

<sup>a</sup> Adjusted hazard ratios and 95% confidence intervals (CI) from Cox regression models with an interaction term between variant (BA.2 vs BA.1). The models were stratified for specimen week, and additionally using regression adjustments for age group, linear effect in age and vaccination status, to account for confounders.

## Discussion

These results suggest that there is no statistically significant difference in the risk of hospital admission between BA.1 and BA.2 cases treated with Sotrovimab in this English community cohort. This absence of difference persists when restricting to those admitted with a diagnosis of COVID-19.

These findings are supported by the access to high quality data on therapeutics prescribed to COVID-19 cases linked to national datasets, permitting a timely nationwide cohort study. Ascertaining case variant status through multiple methods allowed for a greater sample size than would otherwise be possible. Stratifying by specimen week and removing cases with insufficient follow-up time aimed to mitigate the reporting lags observed in routine hospitalisation surveillance data, but some bias may remain.

There are several limitations to this study, primarily NHS numbers were required to link the therapeutic data to the COVID-19 cases data, and subsequent vaccination and hospitalisation data which meant 1,125 community Sotrovimab records were excluded from the analysis. This analysis was also limited by overall number of cases available for inclusion, with variable testing guidance in England throughout the period and restrictions in free community testing from 1 April 2022. Whilst therapeutic agent recipients and hospitalised cases are prioritised for both testing and sequencing (13), the period saw a significant reduction in sequencing capacity which may have limited the study size. Additional limitations include lack of information on co-morbidities of cases, and alternative measures of clinical severity such as people requiring ventilator support or admittance to intensive care units. Furthermore, the vaccination data available did not include the spring booster doses at the time of analysis.

It remains possible that Sotrovimab has lower clinical effectiveness for people with both BA.1 and BA.2, compared to previously circulating variants such as Delta (B.1.617.2), however due to the lack of cases of BA.2 and Delta occurring in the same time period, it is not possible to compare severe outcomes following Sotrovimab treatment between these cases.

While this analysis shows that there is no apparent reduction in Sotrovimab effectiveness against BA.2 compared to BA.1 in this cohort, it is important that Sotrovimab continues to be monitored so that treatment guidance can be updated when necessary. This is particularly important as new variants emerge, such as the BA.4 and BA.5 lineages of Omicron.

## Data Availability

The individual-level nature of the data used risks individuals being identified, or being able to self-identify, if the data are released publicly. Requests for access to these non-publicly available data should be directed to UKHSA.

## Acknowledgments

The authors appreciate the contributions of Tommy Nyberg, Anne Presanis and Daniela De Angelis from MRC Biostatistics Unit, University of Cambridge for their expert advice.

## Author contributions

ST, KH, and GD conceived and designed the study. KH prepared the datasets and performed the statistical analysis, supported by HHW. KH and SGN drafted the first version of the manuscript. All authors read, revised and approved the final version of the manuscript.

## Ethics Approval

UKHSA has legal permission, provided by Regulation 3 of The Health Service (Control of Patient Information) Regulations 2002 to process confidential patient information under Sections 3(i) (a) to (c), 3(i)(d) (i) and (ii) and 3(iii) as part of its outbreak response activities. This study falls within the research activities approved by the UKHSA Research Ethics and Governance Group.

## Consent to Participate

Not applicable.

## Consent to Publish

Not applicable.

## References

1. Medicines and Healthcare products Regulatory Agency. Regulatory approval of Ronapreve 2021 [Available from: https://www.gov.uk/government/publications/regulatory-approval-of-ronapreve.

2. Gupta A, Gonzalez-Rojas Y, Juarez E, Crespo Casal M, Moya J, Rodrigues Falci D, et al. Effect of Sotrovimab on Hospitalization or Death Among High-risk Patients With Mild to Moderate COVID-19: A Randomized Clinical Trial. Jama. 2022;327(13):1236–46.

3. Martin-Blondel G, Marcelin A-G, Soulié C, Kaisaridi S, Lusivika-Nzinga C, Dorival C, et al. Outcome of very high-risk patients treated by Sotrovimab for mild-to-moderate COVID-19 Omicron, a prospective cohort study (the ANRS 0003S COCOPREV study). J Infect. 2022:S0163-4453(22)00196-7.

4. Takashita E, Kinoshita N, Yamayoshi S, Sakai-Tagawa Y, Fujisaki S, Ito M, et al. Efficacy of Antiviral Agents against the SARS-CoV-2 Omicron Subvariant BA.2. New England Journal of Medicine. 2022;386(15):1475–7.

5. Zhou H, Tada T, Dcosta BM, Landau NR. Neutralization of SARS-CoV-2 Omicron BA.2 by Therapeutic Monoclonal Antibodies. bioRxiv. 2022:2022.02.15.480166.

6. U.S. Food & Drug Administration. FDA updates Sotrovimab emergency use authorization 2022 [Available from: https://www.fda.gov/drugs/drug-safety-and-availability/fda-updates-sotrovimab-emergency-use-authorization.

7. Department of Health and Social Care, The Scottish Government, Welsh Government, Department of Health, NHS. Interim Clinical Commissioning Policy: Antivirals or neutralising monoclonal antibodies in the treatment of hospital-onset COVID-19 (Version 6) 2022 [Available from: https://www.england.nhs.uk/coronavirus/publication/interim-clinical-commissioning-policy-antivirals-or-neutralising-monoclonal-antibodies-in-the-treatment-of-hospital-onset-covid-19/.

8. NHS England. COVID-19 Therapeutics (antivirals, neutralising monoclonal antibodies and interleukin 6 inhibitors) 2022 [Available from: https://www.england.nhs.uk/statistics/statistical-work-areas/covid-therapeutics-antivirals-and-neutralising-monoclonal-antibodies/.

9. UK Health Security Agency. COVID-19 therapeutic agents: technical briefing 3 2022 [Available from: https://www.gov.uk/government/publications/covid-19-therapeutic-agents-technical-briefings.

10. Clare T, Twohig KA, O’Connell AM, Dabrera G. Timeliness and completeness of laboratory-based surveillance of COVID-19 cases in England. Public Health. 2021;194:163–6.

11. Twohig KA, Nyberg T, Zaidi A, Thelwall S, Sinnathamby MA, Aliabadi S, et al. Hospital admission and emergency care attendance risk for SARS-CoV-2 delta (B.1.617.2) compared with alpha (B.1.1.7) variants of concern: a cohort study. Lancet Infect Dis. 2022;22(1):35–42.

12. UK Health Security Agency. UK Health Security Agency (UKHSA). SARS-CoV-2 variants of concern and variants under investigation in England. Technical briefing 36 London 2022 [Available from: https://www.gov.uk/government/publications/investigation-of-sars-cov-2-variants-technical-briefings.

13. Cabinet Office. COVID-19 Response: Living with COVID-19 2022 [Available from: https://www.gov.uk/government/publications/covid-19-response-living-with-covid-19.

